# Systematic comparison of quantity and quality of RNA recovered with commercial FFPE tissue extraction kits

**DOI:** 10.1101/2024.03.17.24304077

**Authors:** Sukoluhle Dube, Sharefa Al-Mannai, Li Liu, Sara Tomei, Apryl Sanchez, William Mifsud, Davide Bedognetti, Wouter Hendrickx, Christophe Michel Raynaud

## Abstract

**Background:** FFPE tissue samples are commonly used in biomedical research and are a valuable source for next-generation sequencing in oncology, however, extracting RNA from these samples can be difficult the quantity and quality achieved can impact the downstream analysis. This study compared the effectiveness of seven different commercially available RNA extraction kits specifically designed for use with FFPE samples in terms of the quantity and quality of RNA recovered.

**Methods:** This study used 9 samples of FFPE tissue from three different types of tissue (Tonsil, Appendix and lymph node of B-cell lymphoma) to evaluate RNA extraction methods. Three sections of 20μm of each sample were combined per sample. The slices were distributed in a systematic manner to prevent any biases. Each of the 7 commercially available RNA extraction kits were used according to manufacturer’s instructions, with each sample being tested in triplicate resulting in a total of 189 extractions. The concentration, RNA integrity number (RIN) and DV200 of each extraction was analysed using a LabChip to determine the quantity and quality of the recovered RNA.

**Results:** This study found that despite processing the FFPE samples in the same standardized way, there were disparities in the quantity and quality of RNA recovered across the different tissue types. Additionally, the study found notable differences in the quantity of RNA recovered when using different extraction kits. In terms of quality, three of the kits performed better than the other four in terms of RNA integrity number (RIN) and DV200 values.

**Conclusion:** Though many laboratories have developed their own protocols for specific tissue types, using commercially available kits is still a popular option. Although these kits use similar processes and extraction procedures, the amount and quality of RNA obtained can vary greatly between kits. In this study, among the kits tested, while the Roche kit, provided a nearly systematic better-quality recovery than other kits, the ReliaPrep FFPE Total RNA miniprep from Promega yielded the best ratio of both quantity and quality on the tested tissue samples.

## Introduction

FFPE samples, which are created by preserving biopsies or autopsy samples in formalin and embedding them in paraffin, are one of the most valuable resources for biomedical research. There are over a billion FFPE samples stored in hospitals and tissue banks worldwide [1], [2]. This method of preservation is preferred for a variety of reasons, including that it maintains the original tissue’s morphological features well for diagnostic purposes and is more cost-effective and simpler to process and store than fresh frozen samples [3]. Additionally, while the proteins, DNA and RNA are of better quality in fresh frozen tissue, the long-term preservation of the sample can lead to RNA and protein degradation even at -80C. Therefore, for extensive long storage FFPE samples will preserve better proteins, RNA and DNA than fresh frozen samples [4].

RNA extraction from formalin-fixed paraffin-embedded (FFPE) tissue samples can be challenging as the quality of RNA can get affected by the oxidation, cross-linking, and other chemical modifications occurring during the fixation and preservation process, which can cause extensive damage to the RNA. RNA was first extracted from FFPE samples in 1988 [5], and since then it has been commonly used for real-time polymerase chain reaction (RT-PCR) protocols. With the advancement of next-generation sequencing (NGS), specifically RNA sequencing (RNASeq), it has become a powerful tool for both research and potential clinical use. Despite the greater instability of RNA compared to DNA, many studies have found a high correlation between RNASeq data from fresh frozen and FFPE samples [2], [6], [7]. However, extracting high-quality RNA from FFPE samples remains a challenge, and the low quantity or poor quality of RNA recovered can negatively impact the reliability of RNASeq data.

Recent advances in technology have led to the development of methods that can effectively extract high-quality RNA from FFPE samples. One such method is the use of specific enzymes and buffers that can degrade the crosslinks formed by formalin fixation and release the RNA. Another approach is using laser capture microdissection (LCM) to physically isolate specific regions of the tissue containing the desired RNA before extraction. Additionally, some companies have also developed commercial kits specifically designed for RNA extraction from FFPE samples that have shown promising results. Many manufacturers have developed kits for extracting RNA from FFPE samples, making the process more straightforward and replicable. However, these kits can vary greatly in their efficiency for different tissue types in terms of the quantity and quality of RNA recovered. To evaluate this variation, in this study, we systematically tested the quantity and quality of RNA recovered with seven commercial kits for manual extraction, using identical samples from three different tissue types.

## Material and methods

### Tissue preparation, sectioning, and storage

Samples from three tonsils from individuals with tonsillitis, three appendixes from individuals with appendix inflammation, and three lymph nodes from patients affected by B-cell lymphoma were surgically removed at Sidra Medicine hospital. After surgery, the samples were processed by the Pathology department of Sidra Medicine hospital for diagnostic purposes following their standard operating procedures. Briefly, the samples were cut into approximate 0.5×1×1 cm blocks and incubated in 10% neutral buffered formalin (manufacturer, #cat number) for 18 to 48 hours. The samples were then paraffin-embedded using a Leica EG1150H (Leica) and stored at room temperature not exceeding 25°C. 5 μm slices were made and H7E stained for diagnostic purposes. A picture of the H&E staining of each sample is provided in supplementary figure 1.

**Figure 1:**
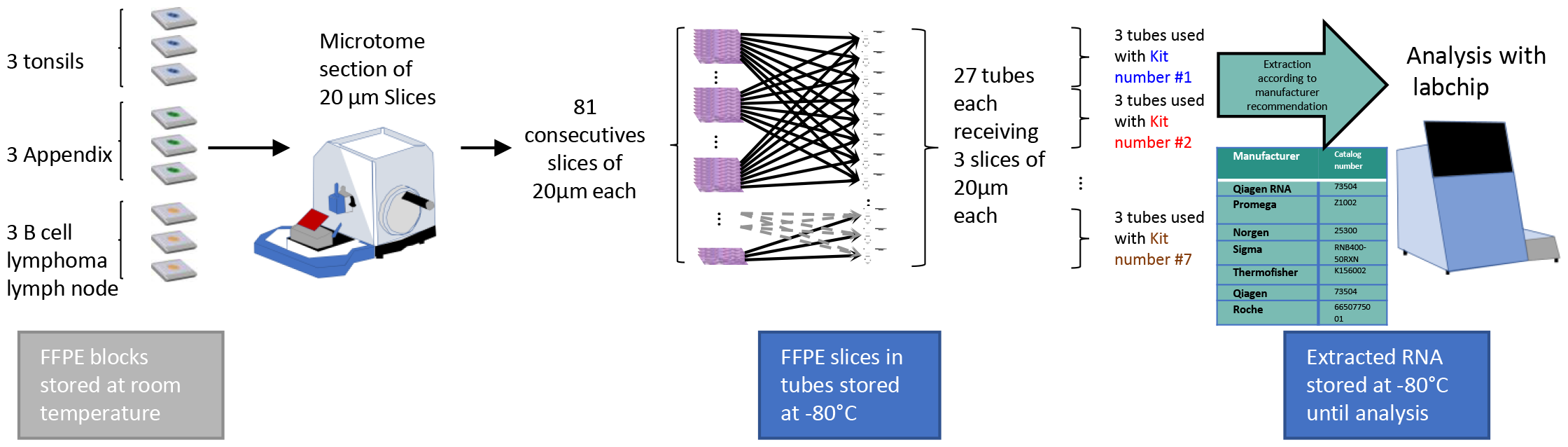
flow chart of tissue sampling. Three independent samples of three types of FFPE tissue (tonsil, appendix and lymph node from B-cell lymphoma) were used in the study, resulting in a total of nine samples. The samples were processed in the pathology lab at Sidra medicine hospital following standard operating procedures. The samples were fixed in formalin for 19-48 hours, embedded in paraffin, and stored at room temperature as FFPE blocks. Each block was then cut into 20 μm sections using a microtome (Leica, #RM2255 Rotary). To avoid regional biases in the tissue sample, a total of 81 sections were cut from each block and distributed systematically across 27 collection tubes. Each tube received three sections of tissue, with one section taken from every other 27 sections. The tubes were then stored at -80°C until extraction with each kit. Each tissue sample was extracted in triplicate using each kit, for a total of 27 extractions per kit and a total of 189 extractions. The extracted RNA was stored at -80°C and quantitative and qualitative analysis were performed simultaneously using the LabChip^®^ GX Touch™ nucleic acid analyser.

After several months of storage at room temperature in the pathology lab archive, the samples were designated as diagnostic surplus and processed for RNA extraction.

To avoid regional biases of cell type or abundance within each block, slices were systematically distributed across the sample collection tubes. Specifically, 20μm thick slices were cut from the block, distributed to 27 tubes with 3 slices per tube, and then stored at -80°C until RNA extraction (less than 1 month). As illustrated in Figure 1, a total of 81 slices of 20μm each were cut and distributed among 27 tubes, with each tube receiving one slice from every 27 cuts and avoiding consecutive slices in the same tube.

### RNA extraction

RNA extraction was performed using each kit listed below according to manufacturer specific recommendations. Each kit extraction was performed in separate days by the same operator.

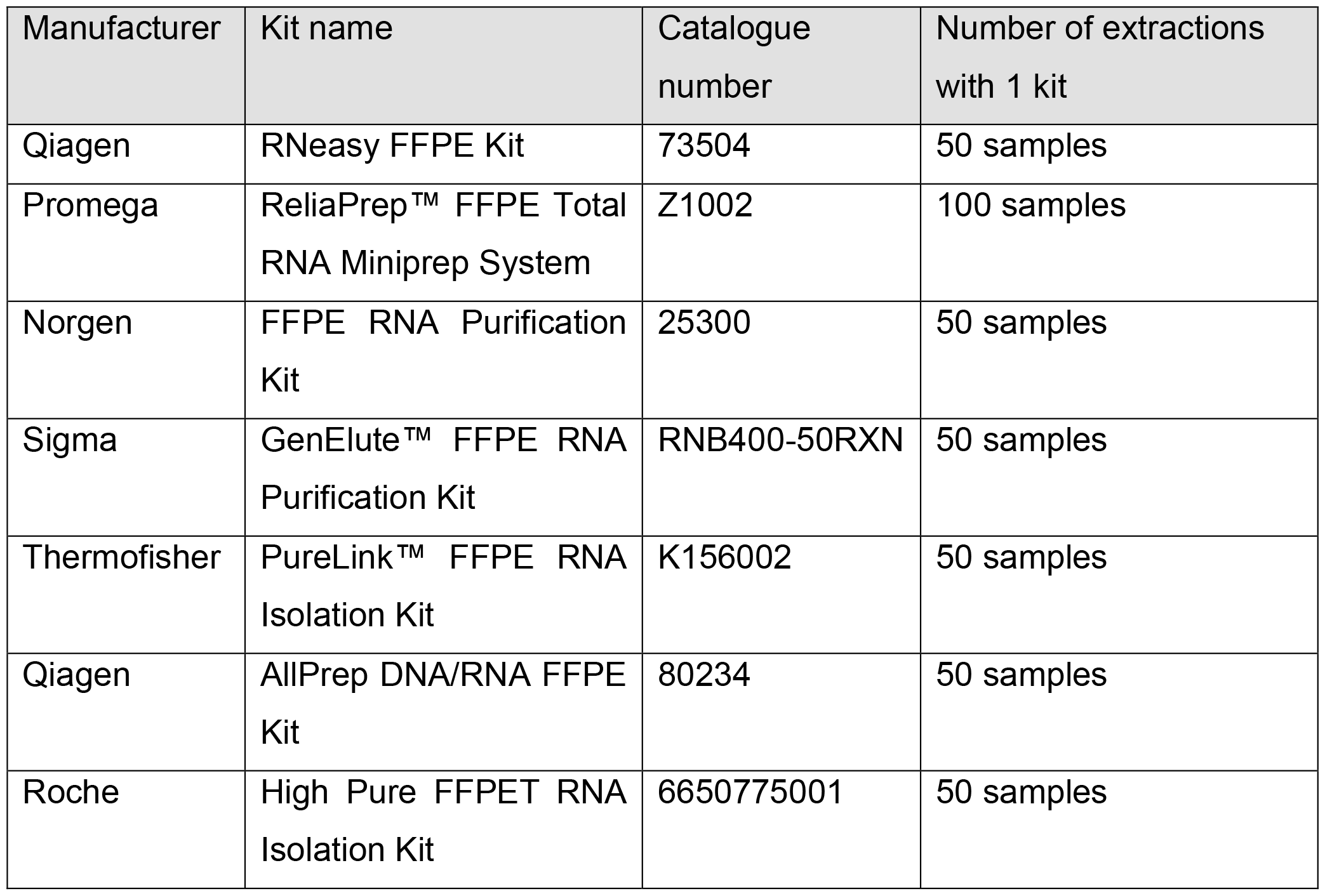

### RNA analysis using LabChip

RNA reagent kits (Catalog# CLS960010) and RNA labchips (Catalog# 760435) were obtained from Perkin Elmer (Waltham, MA). Labchip analysis of sample were performed according to manufacturer specific recommendations. All data obtained are summarised in supplementary table 1.

### Statistics

Two-tailed unpaired t-tests were performed to determine statistical differences between the extractions of the same samples and across samples between kits.

## Results

### Quantitative analysis

Using LabChip measurements, the quantity of RNA recovered with each kit was first analyzed. To account for variability in tissue processing and tissue type that could affect recovery efficiency, the relative quantity was analyzed based on the best-performing kit for each sample. For all 3 tonsil and all 3 lymph node samples from B-cell lymphoma patients and for one appendix sample, the Promega kit provided the maximum recovery, while for two appendix samples, the kit from Thermofisher performed better than the other kits (Figure 1A-C). Overall, the Promega kit allowed significantly higher quantity recovery than the other kits (Figure 2D).

**Figure 2:**
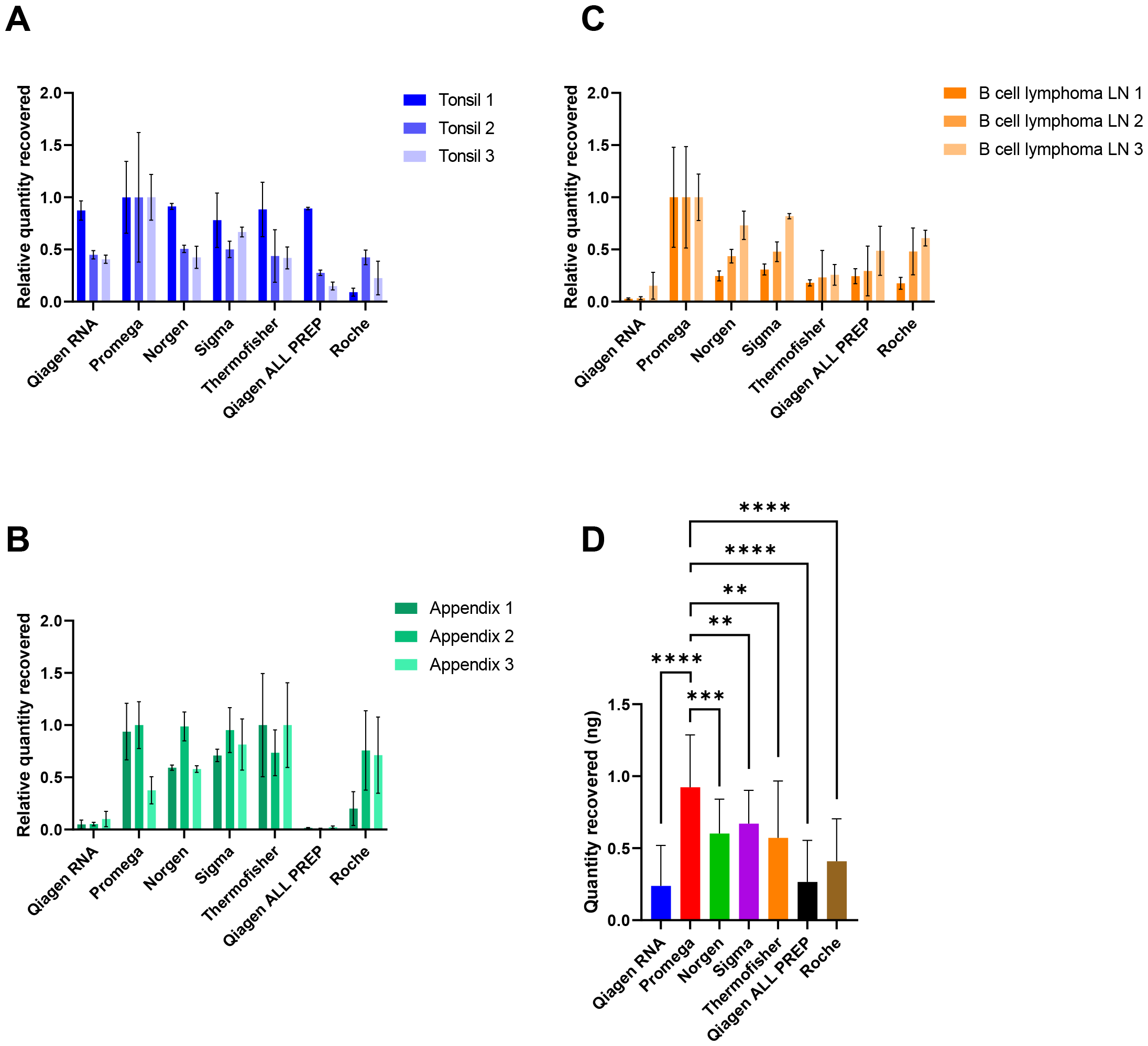
Quantitative comparison of across kits and tissues. (A) Relative quantity of RNA recovered for each tonsil samples compared to best performing kit. (B) Relative quantity of RNA recovered for each appendices samples compared to best performing kit. (C) Relative quantity of RNA recovered for each B cell lymphoma lymph node samples compared to best performing kit. The average and SD of triplicates is represented. N=3 for each bar from the bar chart for figure A-C. (D) Average relative quantity recovered with each kit across the different tissue samples. N=3 for each bar from the bar chart for figure A-C and N=27 for each bar of figure D. P-values were calculated with two tailed unpaired t-test (^**^p<0.01, ^***^p<0.001, ^****^p<0.0001).

### Qualitative analysis

The LabChip system performed two different quality analyses, RIN (on a scale of 1 to 10) and DV200 (expressed in percentage). The RIN (RNA integrity number) is a reliable parameter to assess the integrity of RNA. The RIN value is derived by the size distribution of the RNA and it represents the degree of integrity/degradation of a given sample, with a score of 10 corresponding to intact RNA and a score of 1 corresponding to a highly degraded RNA. The DV200 is a reliable measure of RNA quality representing the percentage of RNA fragments > 200 nt. First, these two quality metrics were analyzed separately. When analyzing RIN values, the Thermofisher kit performed significantly better than the other kits for Tonsils 1 and 2, but for Tonsil 3, Promega, Thermofisher, and Roche kits had no significantly different result, but all three performed better than the other kits (Figure 3A). For Appendixes, Thermofisher and Roche kits performed better than the other kits for appendix 1 and 2, and the Roche kit performed better than any other kit for appendix 3 (Figure 3B). In LN (Lymph Node) samples from B-cell lymphoma, Promega and Roche kits performed significantly better than the other kits except for LN3 where Roche kit performed better than all other kits (Figure 3C).

**Figure 3:**
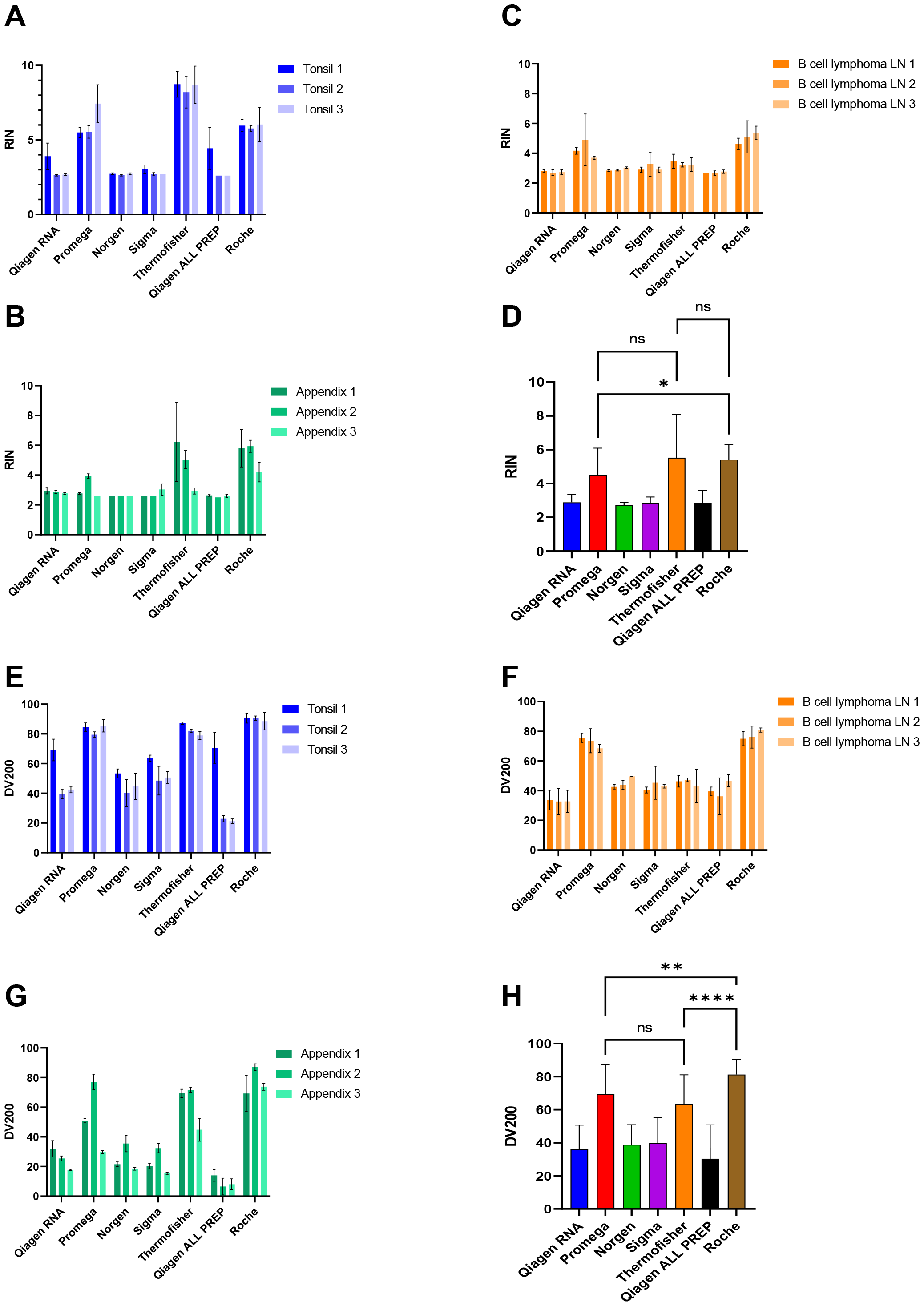
Qualitative comparison of across kits and tissues. (A-C) Quality analysis by RIN of 3 tonsils (A), of 3 appendices (B) and 3 B cell lymphoma lymph nodes (C). (D) Average quality measured by RIN with each kit across the different tissue samples. (E-G) Quality analysis by DV200 of 3 tonsils (E), of 3 appendices (F) and 3 B cell lymphoma lymph nodes (G). (H) Average quality measured by DV200 with each kit across the different tissue samples. N=3 for each bar from the bar chart for figure A-C; E-G and N=27 for each bar of figure D and H. P-values were calculated with two tailed unpaired t-test (^*^p<0.05, ^**^p<0.01, ^****^p<0.0001).

When considering the results altogether, Promega, Thermofisher, and Roche kits performed better in RIN quality analysis across all sample types. However, while the Thermofisher and Roche kits were statistically equivalent, the Roche kit performed slightly but significantly better than the Promega kit (as seen in Figure 3D).

When analyzing the DV200 values, for Tonsils 1 and 3, the kits from Promega, Thermofisher, and Roche performed similarly and better than the other kits, but in Tonsil 2, the Roche kit performed better than any other kit (Figure 3E). For appendixes, Thermofisher and Roche kits performed better than the other kits, but the Roche kit was significantly better than any other kit for appendices 2 and 3 (Figure 3F). Finally for samples from LN from B-cell lymphoma patients, the kits from Promega and Roche performed better than other kits, but Roche was better than any other kit in LN3 (Figure 3G). Overall, Promega, Thermofisher, and Roche kits also performed better in DV200 quality analysis than the other kits across all sample types, but the Roche kit performed significantly better than all of them (Figure 3H).

### Multiparametric analysis

Previously, we found that the Promega kit allows for better recovery of RNA in terms of quantity than its competing kits, and the kits from Promega, Thermofisher, and Roche all allow for better quality of the RNA recovered, with the Roche kit providing the best DV200 values across all samples. When considering RNA sequencing from FFPE samples and depending on the tissue size, some researchers may consider RIN or DV200 more important while others may choose the quantity recovered as the most critical parameter. We therefore represented each of these parameters against each other in figure 4. When RIN and DV200 parameters are taken together we can clearly see that of Thermofisher Promega and Roche constitute a group detached from the rest of the kits (Figure 4A), the individual distribution of the samples shows that while samples recovered with Roche are always in the upper quadrant, some samples isolated with Promega, and even more with Thermofisher, cluster with the rest of the less performant kits (4B). It seems, therefore, that the Thermofisher kit and to a lesser extent, the kit from Promega, are more sensitive to individual variability between samples for quality performance.

**Figure 4:**
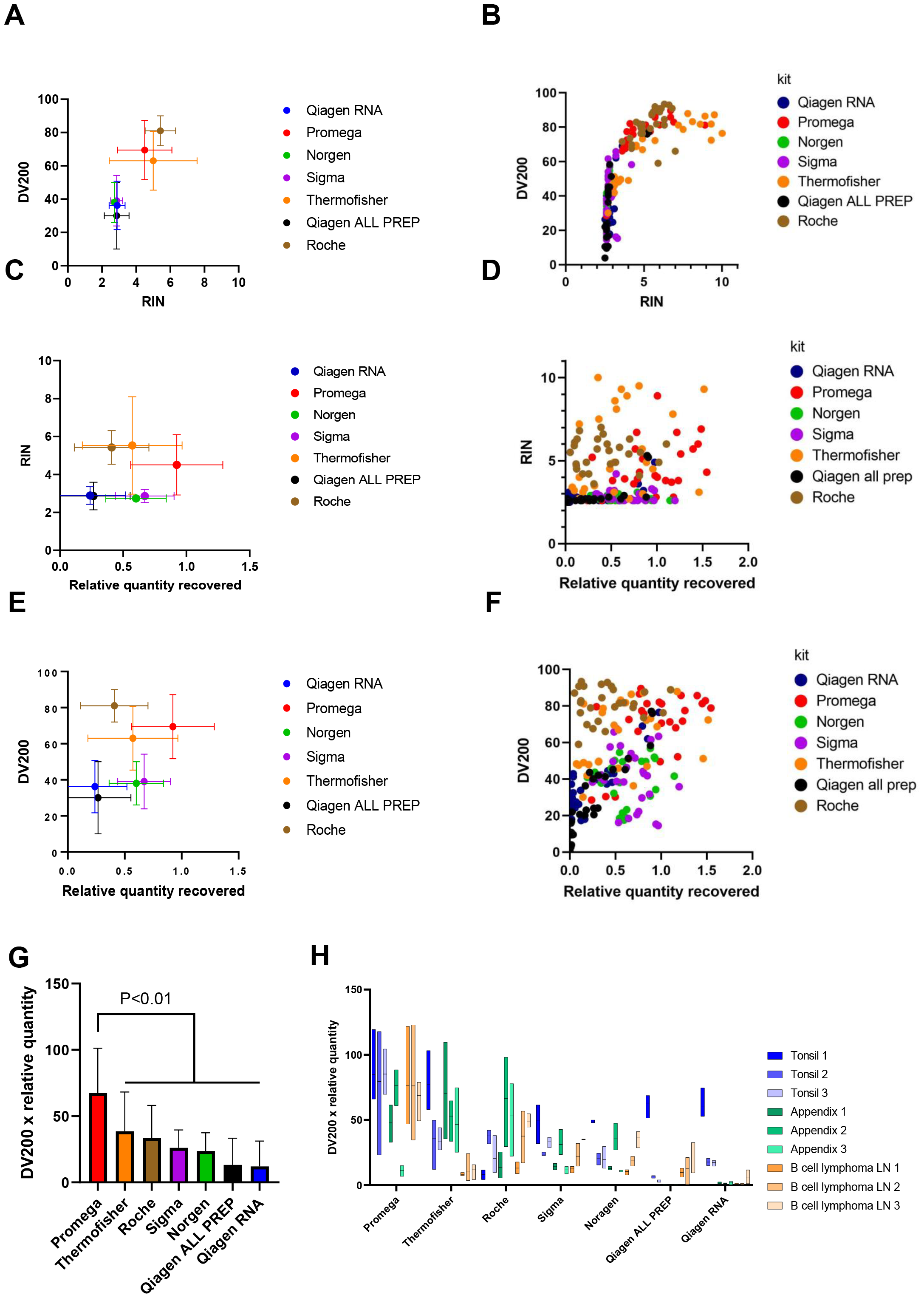
Multiparametric analysis. (A-B) Comparison of quality analysed by RIN and DV200. (A) representation of all samples tested. (B) Average and SD of RIN and DV200 across the different tissue samples. (C-D) Comparison of quality analysed by RIN and average relative quantity recovered with each kit across the different tissue samples. (C) Representation of all samples tested. (D) Average and SD of RIN and average quantity across the different tissue samples. (E-F) Comparison of quality analysed by DV200 and average relative quantity recovered with each kit across the different tissue samples. (E) Representation of all samples tested. (F) Average and SD of DV200 and relative quantity across the different tissue samples. (G) Comparison of score including quality and quantity by multiplying relative quantity recovered by DV200 for each sample. N=27 for each bar of figure G (p<0.01 between Promega and Thermo kits, p<0.0001 between Promega and all other kits). (H) Box plot of relative quantity x DV200 score for each sample.

When RIN or DV200 and the average quantity recovered are considered together, while on average, the kits from Promega, Thermofisher and Roche again out-perform the other kits (Figure 4C, 4E), only samples isolated with the Roche kit are systematically better than the other kits. The Thermofisher kit can allow exceptionally great quality recovery in some samples but largely lower quality in others. Similarly, but to a lesser extent, with the Promega kit, some samples’ quality can be lower (Figure 4D, 4F).

To identify the kit that offered the best compromise between quantity and quality recovered, we created a score by multiplying the relative quantity recovered by DV200 (offering the best dynamic range of analysis between DV200 and RIN). Using this score, we show that Promega significantly outperforms all other kits (Figure 4G), despite presenting a lower score than other kits in some particular samples (Figure 4H).

## Conclusion

When extracting RNA from FFPE samples for RNA sequencing, multiple parameters must be taken into consideration, including the quantity and quality of RNA recovered to allow proper sequencing and coverage for correct data analysis. In the study presented, some kits were systematically outperformed by others, both in terms of quality and quantity recovered. Among them, the historically commonly used kit from Qiagen (both RNA-specific and ALL PREP) was a surprise to us.

In terms of quantity recovered, the Promega kit performed better on average than any of the other kits, but this might come at the expense of quality of the recovered RNA in some samples. Therefore, in cases of small biopsies or small samples, this kit might be the best choice to maximize the recovery rate. However, it is important to note that in some cases, we observed large standard deviations in the quantity recovered between triplicates of the same sample (including with Promega). Our hypothesis is that in some cases, the column used can be slightly clogged, resulting in less volume recovered after final centrifugation and therefore, higher concentration. This would not affect the quality of the RNA recovered, explaining why less variation was observed in terms of RIN and DV200. On the other hand, if sample size is not an issue, and the choice is made to maximize the quality of the sample, one might consider using the Roche kit, which provided a nearly systematic better-quality recovery than other kits.

When trying to identify the best compromise between quantity recovered and quality recovered, a score was created, taking into account both quantity recovered and quality (as measured by DV200). In that analysis, it was observed that Promega gave a significantly better compromise than other kits.

In any case, it is clear that the quality and quantity of RNA recovered can vary greatly between tissue types and samples, likely due to factors such as protein content, fixation parameters, or cell types present within the sample. The standard practice of tissue fixation can vary considerably between institutions and laboratories, and it is important to evaluate the impact of fixation time on the quality and quantity recovered with various kits. Additionally, the thickness of the section used can also affect the results. Some studies have shown that sections with a thickness of 10μm or more perform better than finer sections, but too thick sections might give opposite results. Therefore, in specific settings, some kits that have been shown to perform not so well in this study, may perform better. To get the best results for a specific type of tissue and sample, it is advisable to test a variety of kits. Finally, some companies offer automated systems that employ similar kits, which can be useful when analysing larger cohorts or ensuring better reproducibility.

## Data Availability

The datasets generated and/or analyzed during the current study are available in the supplementary table 1.

## Statements

### Declarations

Ethics approval and consent to participate

“Not applicable”

### Consent for publication

“Not applicable”

## Acknowledgements

The authors would like to acknowledge the Sidra Medicine research branch core facilities and our research administration team without whom we could not have performed this work.

## Author’s contribution

W.M. selected all the samples, S.D. cut all the samples, S.A., S.D., C.R., A.S. performed the experiments, L.L., S.T. performed the RIN and DV200 analysis, C.R., D.B., performed the analysis C.R., drafted the manuscript and the figures, W.H., edited the text. Conceptualization by W.M., C.R., D.B. and W.H., project supervision and coordination C.R. All authors reviewed the manuscript.

---

*Supplementary table 1: raw data of each sample*

The raw data of RIN, DV200, concentration and UV-VIS reading of each samples are provided in this table

